# Neuropsychiatric and cognitive symptoms across the Alzheimer’s disease clinical spectrum: Cross-sectional and longitudinal associations

**DOI:** 10.1101/2021.02.18.21251064

**Authors:** Willem S. Eikelboom, Esther van den Berg, Ellen Singleton, Sara J. Baart, Michiel Coesmans, Annebet E. Leeuwis, Charlotte E. Teunissen, Bart N.M. van Berckel, Yolande A.L. Pijnenburg, Philip Scheltens, Wiesje M. van der Flier, Rik Ossenkoppele, Janne M. Papma

**Affiliations:** Department of Neurology, Erasmus MC, University Medical Center, Rotterdam, the Netherlands; Department of Neurology, Alzheimer Center Amsterdam, Amsterdam University Medical Centers, Amsterdam, the Netherlands; Department of Biostatistics, Erasmus MC, University Medical Center, Rotterdam, the Netherlands; Department of Psychiatry, Erasmus MC, University Medical Center, Rotterdam, the Netherlands; Neurochemistry Laboratory, Department of Clinical Chemistry, Amsterdam Neuroscience, Amsterdam University Medical Centers, Vrije Universiteit, Amsterdam, the Netherlands; Department of Radiology and Nuclear Medicine, Amsterdam University Medical Centers, Amsterdam, the Netherlands; Clinical Memory Research Unit, Lund University, Malmö, Sweden

## Abstract

**Objective:** To investigate the prevalence and trajectories of neuropsychiatric symptoms (NPS) in relation to cognitive functioning in a cohort of amyloid-*β* positive individuals across the Alzheimer’s disease (AD) clinical spectrum.

**Methods:** We included 1,524 amyloid-*β* positive individuals from the Amsterdam Dementia Cohort with subjective cognitive decline (SCD, n=113), mild cognitive impairment (MCI, n=321), or dementia (n=1,090). We measured NPS with the neuropsychiatric inventory (NPI), examining total scores and the presence of specific NPI-items. Cognition was assessed across five cognitive domains and with the MMSE. We examined trajectories including model based trends for NPS and cognitive functioning over time. We used linear mixed models to relate baseline NPI scores to cognitive functioning at baseline (whole-sample) and longitudinal time-points (subsample n=520, Mean=1.8 [SD=0.7] years follow-up).

**Results:** NPS were prevalent across all clinical AD stages (NPI total score ≥1 81.4% in SCD, 81.2% in MCI, 88.7% in dementia). Cognitive functioning showed an uniform gradual decline; while in contrast, large intra-individual heterogeneity of NPS was observed over time across all groups. At baseline, we found associations between NPS and cognition in dementia that were most pronounced for NPI total scores and MMSE (range *β*:-0.18–0.11, FDR-adjusted *p*<0.05), while there were no cross-sectional relationships in SCD and MCI (*β*:-0.32– 0.36, FDR-adjusted *p*>0.05). There were no associations between baseline NPS and cognitive functioning over time in any clinical stage (*β*:-0.13–0.44, FDR-adjusted *p*>0.05).

**Conclusion:** NPS and cognitive symptoms are both prevalent across the AD continuum, but show a different evolution during the course of the disease.

## INTRODUCTION

Alzheimer’s disease (AD) is characterized by a gradual decline in cognitive functions and activities of daily living.^1^ As neuropsychiatric symptoms (NPS) are present in the majority of patients with AD dementia,^2^ NPS are increasingly recognized as core clinical AD symptoms.^3^ Previous studies have associated the presence of NPS with an increased risk of progression to dementia and with worse cognitive performance and a faster cognitive decline in AD dementia.^4-6^ These studies have emphasized the clinical relevance of NPS in AD by highlighting its prognostic value.

Yet, several other studies have not found an association between NPS and cognitive functioning in AD dementia.^7-9^ These discrepant results may have a number of causes, such as the fact that studies often used instruments that assess general cognitive functioning (i.e. Mini-Mental State Examination [MMSE]) and overall NPS burden (i.e. Neuropsychiatric Inventory [NPI] total score).^7, 10^ Furthermore, prior studies have often included patients based on clinical diagnostic criteria of AD without biomarker evidence,^9, 11^ thereby increasing the likelihood of including patients with non-AD primary etiologies.^12^

To address these challenges, the current study investigates 1) the prevalence and course of specific NPS, and 2) associations between baseline NPS and performance on multiple cognitive domains at baseline and over time in an amyloid-*β* positive sample ranging from normal cognition to dementia. This knowledge will provide a better understanding of the manifestation of NPS across the clinical stages of AD and its relationship with cognitive decline which could aid patient management in clinical practice.

## METHODS

### Participants

We included all patients who visited Alzheimer Center Amsterdam between June 2002 and December 2017 and had 1) a clinical diagnosis of subjective cognitive decline (SCD), mild cognitive impairment (MCI), or probable AD dementia, 2) were amyloid-*β* positive, and 3) had NPI and neuropsychological assessment available at baseline. All individuals underwent a standard diagnostic work-up including medical history taking, neurological examination, cognitive testing, lumbar puncture, and brain MRI.^13^ A subsample of the individuals underwent amyloid-*β* PET for research purposes (n=450). Clinical diagnoses were established using conventional diagnostic criteria at multidisciplinary meetings. Individuals had to meet the clinical criteria of SCD,^14^ MCI,^15^ or probable AD dementia,^1^ in addition to amyloid-*β* positivity based on either CSF (i.e. amyloid-*β*_42_ <550 pg/mL or tau/ amyloid-*β*_42_ ratio >0.52)^16^ or visual rating of an amyloid-*β* PET scan with the radiotracers ^18^F-florbetaben (n=190), ^11^CPIB (n=133), ^18^F-flutemetamol (n=100), or ^18^F-florbetapir (n=27).^17^ In case of amyloid-*β* PET/CSF discordance, amyloid-*β* status was determined based on the visual rating of amyloid-*β* PET. This resulted in a total of 1,524 individuals of which 113 participants had a clinical diagnosis of SCD, 321 participants with MCI, and 1,090 participants with AD dementia at baseline. As all participants were amyloid-*β* positive, SCD will be denoted as preclinical AD and MCI as MCI due to AD.

A subsample of the participants had follow-up assessments available: n=53 (46.9%) with preclinical AD at baseline, n=142 (44.2%) with MCI due to AD at baseline, and n=325 (29.8%) with AD dementia at baseline. We conducted longitudinal analyses in patients who had follow-up assessments available limited up to three years after baseline assessment, since less than ten percent of the 1,524 participants had more than 3 years of follow-up assessments available. Including these assessments may have resulted in underestimation of disease progression due to selective drop-out.^18^ For those with follow-up assessment available, mean follow-up duration was 1.7 years (SD=0.8) for preclinical AD, 1.9 years (SD=0.7) for MCI due to AD, and 1.7 years (SD=0.7) for AD dementia.

### Standard Protocol Approvals, Registrations, and Patient Consent

The Medical Ethics Review Committee of the Amsterdam University Medical Centers approved the study. Written informed consent was obtained from all participants.

### Neuropsychiatric assessment

The neuropsychiatric inventory (NPI) was used to assess NPS.^19^ This 12-item informant-based interview is a widely accepted measure of NPS in dementia.^3^ Each item is rated according to its severity (0-3) and frequency (0-4). We multiplied the severity and frequency scores for each item (0-12). The presence of specific NPS was defined as a severity × frequency score of ≥1 for each NPI-item. We summed the severity × frequency scores of all 12 NPI-items to obtain the NPI total score (0-144). The presence of any NPS was defined as a NPI total score of ≥1. At baseline, scores were missing for the following NPI-items: n=8 for eating behaviors, n=8 for night-time behaviors, n=2 for aberrant motor behaviors, n=1 for apathy, and n=1 for agitation.

### Neuropsychological assessment

We used the Mini-Mental State Examination (MMSE) to assess global cognitive functioning. In addition, a standardized neuropsychological test battery was used to measure performance across five cognitive domains. We used immediate recall scores of the Visual Association Test part A and the immediate recall and delayed recall of the Rey Auditory Verbal Learning Test to measure memory. For attention, the Digit Span forward, Stroop Word Color test color and word conditions, and the Trail Making Test part A were administered. Executive functioning was assessed using Digit Span backward, Stroop test color-word condition, Trail Making Test part B, and the Frontal Assessment Battery. We used category fluency (animals) and the naming condition of the Visual Association Test to measure language. We measured visuospatial abilities using the number location, dot counting, and fragmented letters subtests of the Visual Object and Space Perception Battery.

Individuals who were not able to complete the Trail Making Test or Stroop test due to cognitive difficulties were assigned the minimum score. We converted raw test scores into Z scores based on the mean (M) and standard deviation (SD) of an independent healthy reference group of 533 AD-biomarker negative individuals (M[SD] age=59.7[9.8], 54% female, M[SD] MMSE score=28.9[1.0]).^20^ The Z scores of the Trail Making Test and Stroop test were inverted to ensure that lower scores indicated worse performance. Next, Z scores were combined into cognitive domain scores by averaging cognitive scores if at least two tests within that domain were available for that individual. At baseline, cognitive domain scores were missing for 7 to 28%.

### Statistical analysis

We compared baseline clinical characteristics, NPS prevalence, and cognitive performance across the diagnostic groups using ANOVA (with Tukey’s HSD post hoc test) or Chi-squared tests where appropriate.

We aimed to statistically analyze trajectories of NPI-scores; yet, the assumption of normality was not met for the longitudinal NPI-item scores given the substantial proportion of zeros, which remained unchanged after deploying several transformations. Therefore, we plotted individual trajectories of NPS and cognitive functioning over time according to disease stage and added model based trends with 95% confidence intervals to the graphs for descriptive purposes. In addition, we investigated the extent to which NPS and cognitive functioning changed over time within individuals (intra-individual variance) and between individuals (inter-individual variance). To quantify the variation within and between individuals, we conducted multilevel null models to obtain the percentage variance explained by intra-variance and inter-variance for neuropsychiatric measures and cognitive measures over time. For these analyses, the continuous severity x frequency scores (0-12) of specific NPI-items were used.

To study associations between baseline NPS and cognitive functioning at baseline and over time, we performed linear mixed models (LMMs) including random intercepts and fixed slopes that were corrected for age, sex, and education. Determinants included the NPI total score and the presence of specific NPI-items with a focus on the three most prevalent NPI-items based on the aforementioned analysis. Outcomes were performance on the MMSE and the five predefined cognitive domains. LMMs were run separately for the clinical stages at baseline (i.e. preclinical AD, MCI due to AD, AD dementia). We tested non-linear associations using LMMs with quadratic and cubic splines and selected linear LMM for all models based on the Likelihood-Ratio Chi-Squared Test and Akaike information criterion.

We checked assumptions by visual inspection of standardized residuals scatterplots and Q-Q plots. As normality of cognitive scores slightly deviated in language and visuospatial abilities most pronounced in preclinical AD, we conducted sensitivity analyses using a bootstrap procedure with 200 bootstrap samples to calculate confidence intervals. This approach did not change the initial findings.

Level of significance was set at *p*<0.05. Yet, the post-hoc analyses on the NPS prevalence rates and the LMMs to study associations between NPS and cognitive performance were corrected for multiple testing using the Benjamin-Hochberg adjusted false discovery rate (FDR) of 0.05. Analyses were performed using *SPSS* version 26.0 and *R* version 4.0 (*lme4, splines, lmerTest, effectsize*, and *boot* packages).

### Data Availability

Data not provided in the article and additional information on methods and materials can be shared upon reasonable request.

## RESULTS

### Participants

Baseline demographic and clinical characteristics of our sample of amyloid-*β* positive individuals are shown in Table 1. Participants with MCI due to AD were older than individuals with AD dementia (*p*<0.001) or SCD (*p*<0.05). Participants with AD dementia had lower levels of education compared to those without dementia (*p*<0.001). The proportion of females was higher in AD dementia than MCI due to AD (*p*<0.001). Of the individuals without dementia at baseline who had follow-up assessment available, 24.5% (n=13) of the individuals with preclinical AD progressed to MCI or dementia, and 43.0% (n=61) of the participants with MCI due to AD progressed to dementia. As expected, baseline MMSE and baseline cognitive domain scores differed according to disease stage (*p*<0.001; table 1).

**Table 1.** Demographic and clinical characteristics of the amyloid-*β* positive sample at baseline according to clinical AD stage.

|  | **Preclinical AD (n = 113)** | **MCI due to AD (n = 321)** | **AD dementia (n = 1,090)** |
| --- | --- | --- | --- |
| **Age, mean ± SD** | 65.8 ± 7.4 | 67.7 ± 7.3 | 65.9 ± 7.7<sup>b</sup> |
| **Female, n (%)** | 52 (46.0%) | 135 (42.1%) | 571 (52.4%)<sup>b</sup> |
| **Education level<sup>a</sup>, median [IQR]** | 6 [1] | 5 [2] | 5 [2]<sup>c</sup> |
| **MMSE score, mean ± SD** | 28.0 ± 1.7 | 26.3 ± 2.4<sup>f</sup> | 20.3 ± 5.1<sup>c</sup> |
| **Cognitive Z scores, mean ± SD** |  |  |  |
| **Memory** | -0.39 ± 0.9 | -2.40 ± 1.7<sup>f</sup> | -4.31 ± 2.1<sup>c</sup> |
| **Attention** | -0.26 ± 0.9 | -0.68 ± 1.2 | -3.50 ± 4.0<sup>c</sup> |
| **Executive functioning** | -0.22 ± 0.9 | -0.80 ± 1.0<sup>f</sup> | -2.68 ± 1.7<sup>c</sup> |
| **Language** | -0.11 ± 0.6 | -0.55 ± 0.7<sup>g</sup> | -1.68 ± 1.6<sup>b</sup> |
| **Visuospatial abilities** | -0.08 ± 0.7 | -0.39 ± 0.9 | -2.97 ± 3.8<sup>c</sup> |
| **NPI total score, mean ± SD** | 8.1 ± 9.2 | 7.8 ± 9.0 | 10.9 ± 10.6<sup>d</sup> |
| **No. of NPS present, mean ± SD** | 2.4 ± 1.9 | 2.4 ± 1.9 | 2.9 ± 2.1<sup>b</sup> |
| **Clinical FU available, n (%)** | 53 (46.9%) | 142 (44.2%) | 325 (29.8%) |
| **No. FU assessments available, mean ± SD** | 1.7 ± 0.8 | 1.7 ± 0.7 | 1.6 ± 0.7 |
| **FU duration in years, mean ± SD** | 1.7 ± 0.8 | 1.9 ± 0.7 | 1.7 ± 0.7 |
| **Progressed to MCI or dementia, n (%)** | 13 (24.5%) | 61 (43.0%) | N/A |

Abbreviations: AD = Alzheimer's disease, MCI = mild cognitive impairment, NPI = neuropsychiatric inventory, FU = follow-up, MMSE = Mini-Mental State Examination.
<sup>a</sup> Dutch education system categorized into levels ranging from 1= less than six years of primary education to 7 = academic degree.
<sup>b</sup> AD dementia < MCI due to AD, $p < 0.001$ .
<sup>c</sup> AD dementia < MCI due to AD; AD dementia < preclinical AD, $p < 0.001$ .
<sup>d</sup> AD dementia < MCI due to AD; AD dementia < preclinical AD, $p < 0.05$ .
<sup>e</sup> MCI due to AD < preclinical AD, $p < 0.001$ .
<sup>f</sup> MCI due to AD < preclinical AD, $p < 0.01$ .
<sup>g</sup> MCI due to AD < preclinical AD, $p < 0.05$ .

### Prevalence of NPS at baseline across clinical stages

NPS were prevalent across all AD stages with at least one NPS present in 81.4% of the individuals with preclinical AD, 81.2% of the individuals with MCI due to AD, and 88.7% of the individuals with AD dementia. The NPI total score was higher for AD dementia compared to preclinical AD and MCI due to AD (*p*<0.05), while we found no difference in NPI total score between preclinical AD and MCI due to AD (*p*=0.97, table 1). The number of NPS present at baseline was higher for AD dementia compared to MCI due to AD (*p*<0.001), with no difference between preclinical AD and the other clinical stages (all *p*>0.05, table 1). The prevalence rates of the specific NPI-items across the clinical AD stages are presented in figure 1. The three most prevalent NPS were similar for all clinical stages and included apathy, irritability, and depression. The prevalence was higher at the more advanced clinical stage for the majority of NPI-items, especially for apathy, anxiety, eating behaviors, aberrant motor behaviors, and delusions. However, irritability, depression, nighttime behaviors, and hallucinations were more common in preclinical AD compared to MCI due to AD and/or AD dementia. The NPI severity scores and frequency scores showed a similar pattern as the NPI prevalence rates, i.e. the highest severity and frequency scores were seen in AD dementia with little differences between preclinical AD and MCI due to AD (table e-1).

**Figure 1.**
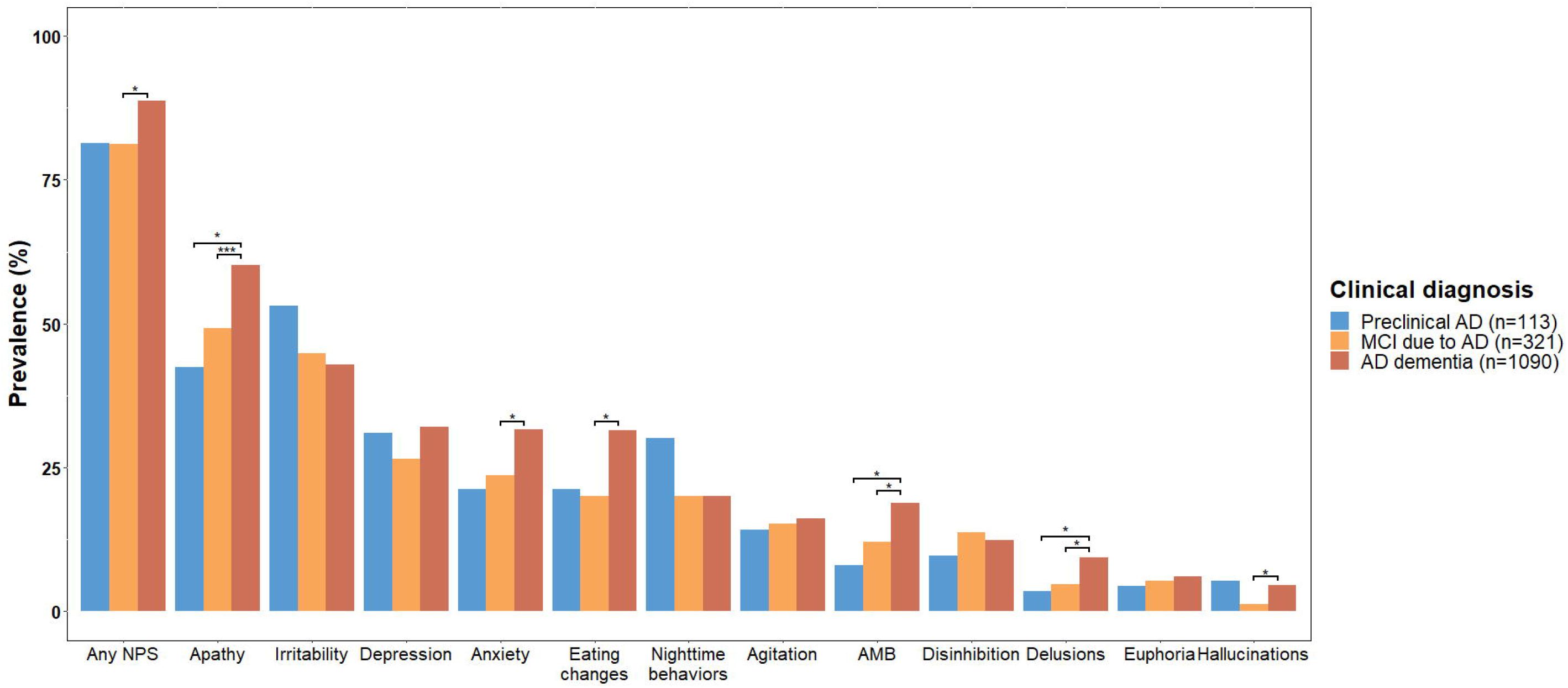
Prevalence of neuropsychiatric symptoms across an amyloid-*β* positive sample according to clinical AD stage. Abbreviations: AD = Alzheimer’s disease; MCI = mild cognitive impairment, NPS = neuropsychiatric symptoms, AMB = aberrant motor behaviors. * *p*<0.05, ** *p*<0.01, *** *p*< 0.001, after correcting for false discovery rate.

### Progression of NPS and cognition over time across clinical stages

We plotted trajectories of specific NPI-items and performance on specific cognitive domains over time for patients across the different clinical AD stages. In participants with preclinical AD at baseline, the trends of specific NPI-item scores remained stable over time with a decline in apathy and a subtle increase for depression, anxiety, and agitation (figure 2). Cognitive scores remained relatively stable over time for preclinical AD. In participants with MCI due to AD at baseline, we observed a relatively stable trends of specific NPI-items over time, whereas a decline was observed in all cognitive domains (figure 3). In participants with AD dementia, little changes were found in trends of specific NPI-items over time, with modest increases in irritability, aberrant motor behaviors, and nighttime behaviors and decrease in depression and anxiety (figure 4). Substantial decline was observed in all cognitive domains.

**Figure 2.**
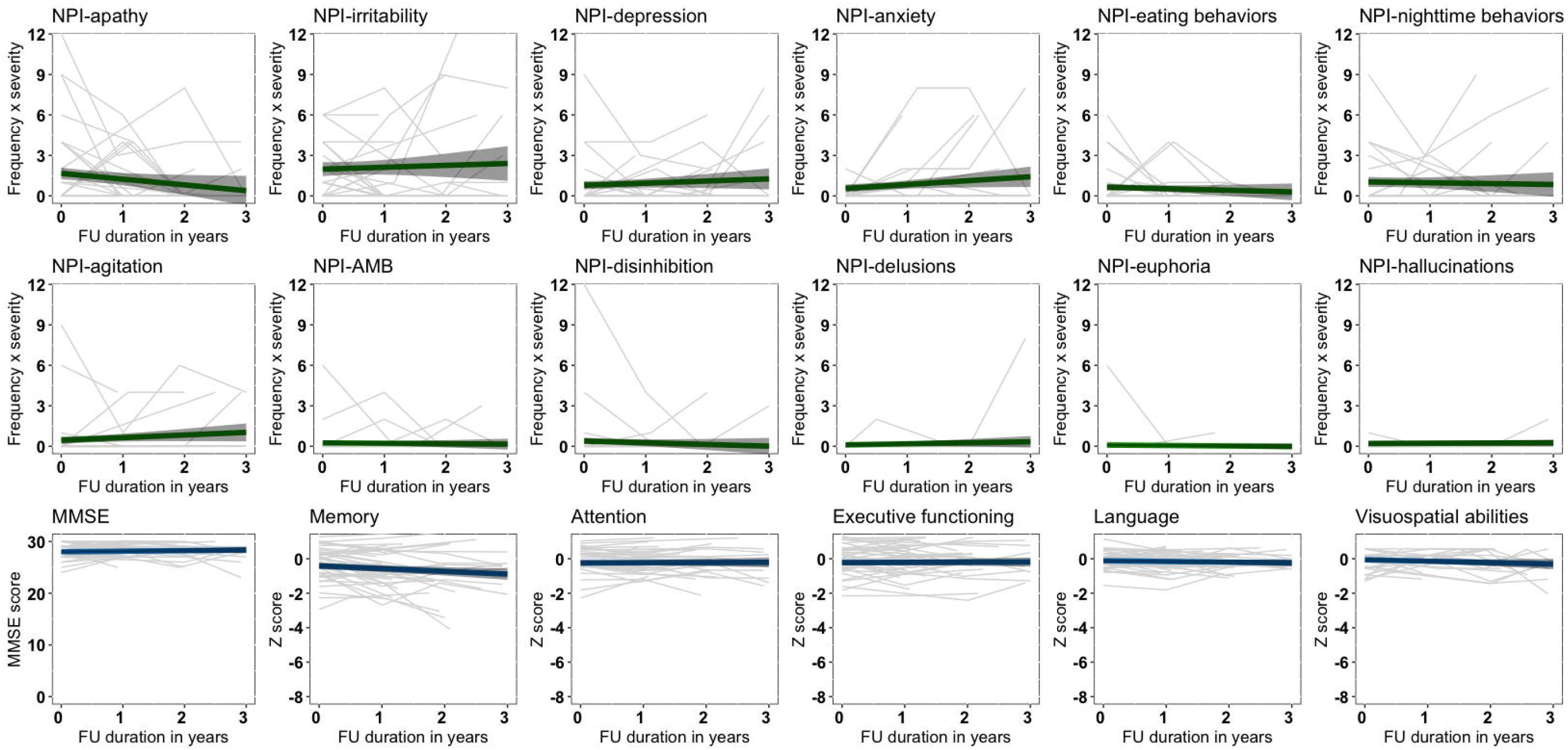
Longitudinal NPI-scores and cognitive functioning for individuals with preclinical AD at baseline. Abbreviations: NPI = Neuropsychiatric Inventory, AD = Alzheimer’s disease, MMSE = Mini-Mental State Examination, AMB = aberrant motor behaviors. Individual trajectories are depicted with model based trends with 95% confidence intervals.

**Figure 3.**
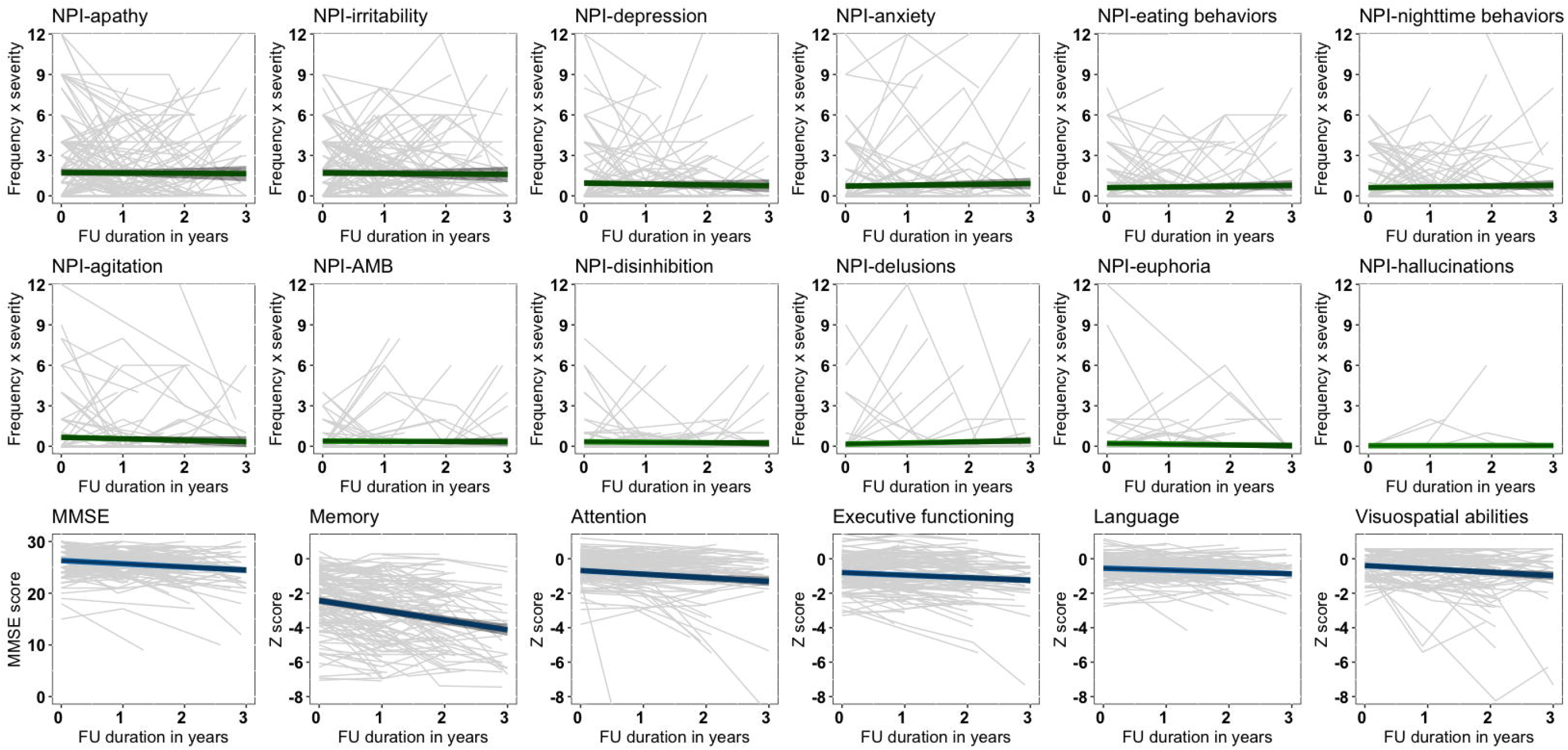
Longitudinal NPI-scores and cognitive functioning for patients with MCI due to AD at baseline. Abbreviations: NPI = Neuropsychiatric Inventory, MCI = mild cognitive impairment, AD = Alzheimer’s disease; MMSE = Mini-Mental State Examination, AMB = aberrant motor behaviors. Individual trajectories are depicted with model based trends with 95% confidence intervals.

**Figure 4.**
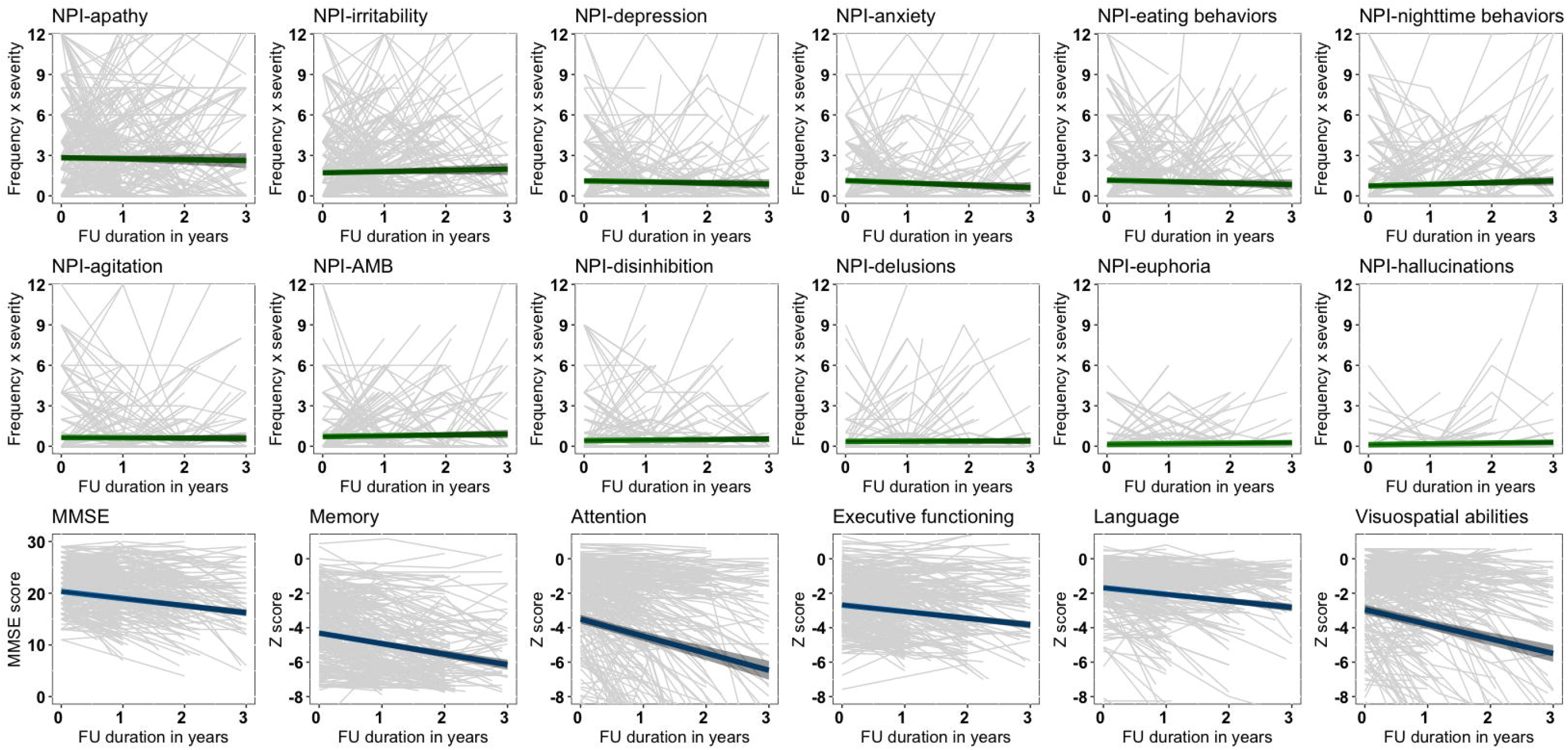
Longitudinal NPI-scores and cognitive functioning for patients with AD dementia at baseline. Abbreviations: NPI = Neuropsychiatric Inventory, AD = Alzheimer’s disease, MMSE = Mini-Mental State Examination, AMB = aberrant motor behaviors. Individual trajectories are depicted with model based trends with 95% confidence intervals.

When looking at the trajectories of specific NPS and cognitive scores over time, we observed large variability in the course of specific NPS within and between individuals across all clinical stages (figures 2-4). To further quantify this intra-individual variability and inter-individual variability, we performed multilevel null models for each measure according to clinical stage at baseline (table e-2). Across all clinical AD stages, the intra-individual variance of NPS measures was higher (all mean % explained >70%) compared to cognitive measures (all mean % explained <45%). Hence, we observed larger changes on NPS measures over time within individuals than between individuals, while the opposite was the case for cognitive measures.

### Cross-sectional associations between NPS and cognitive functioning at baseline

Age-, sex-, and education-corrected LMM in AD dementia showed that higher baseline NPI total scores were associated with lower baseline MMSE scores (FDR-adjusted *p*<0.001) and lower performance on visuospatial abilities (FDR-adjusted *p*<0.05). Baseline NPI total scores were not related to cognitive functioning at baseline in preclinical AD and MCI due to AD (all FDR-adjusted *p*>0.05, table 2).

**Table 2.** Associations between baseline NPS and concurrent cognitive performance according to clinical AD stage.

| **Preclinical AD (n=113)** |  |  |  |  |  |  |
| --- | --- | --- | --- | --- | --- | --- |
|  | **MMSE** | **Memory** | **Attention** | **EF** | **Language** | **Visuospatial** |
| **NPI total** | -0.07 [-0.27, 0.12] | 0.00 [-0.21, 0.21] | -0.06 [-0.27, 0.14] | -0.20 [-0.39, -0.01] | -0.13 [-0.34, 0.09] | 0.13 [-0.07, 0.32] |
| **Apathy** | -0.20 [-0.56, 0.16] | -0.19 [-0.58, 0.20] | 0.25 [-0.12, 0.63] | 0.17 [-0.19, 0.53] | 0.00 [-0.40, 0.40] | -0.01 [-0.39, 0.38] |
| **Depression** | -0.12 [-0.46, 0.23] | -0.13 [-0.50, 0.24] | -0.24 [-0.59, 0.12] | -0.02 [-0.36, 0.33] | -0.23 [-0.61, 0.14] | 0.28 [-0.06, 0.62] |
| **Irritability** | 0.12 [-0.22, 0.46] | 0.36 [0.00, 0.72] | 0.14 [-0.21, 0.50] | -0.09 [-0.43, 0.24] | -0.32 [-0.68, 0.05] | 0.19 [-0.15, 0.53] |
| **MCI due to AD (n=321)** |  |  |  |  |  |  |
|  | **MMSE** | **Memory** | **Attention** | **EF** | **Language** | **Visuospatial** |
| **NPI total** | -0.03 [-0.13, 0.06] | 0.04 [-0.06, 0.14] | -0.07 [-0.17, 0.03] | -0.07 [-0.17, 0.03] | -0.06 [-0.17, 0.05] | -0.02 [-0.13, 0.09] |
| **Apathy** | -0.08 [-0.27, 0.11] | 0.06 [-0.15, 0.26] | -0.17 [-0.37, 0.04] | -0.05 [-0.25, 0.15] | -0.10 [-0.32, 0.13] | -0.04 [-0.26, 0.18] |
| **Depression** | -0.15 [-0.36, 0.07] | -0.16 [-0.39, 0.07] | -0.18 [-0.41, 0.05] | -0.09 [-0.31, 0.12] | -0.25 [-0.49, 0.00] | -0.04 [-0.28, 0.21] |
| **Irritability** | 0.05 [-0.14, 0.24] | 0.10 [-0.11, 0.30] | -0.13 [-0.33, 0.08] | -0.07 [-0.27, 0.12] | -0.04 [-0.26, 0.18] | 0.03 [-0.19, 0.25] |
| **AD dementia (n=1,090)** |  |  |  |  |  |  |
|  | **MMSE** | **Memory** | **Attention** | **EF** | **Language** | **Visuospatial** |
| **NPI total** | **-0.08 [-0.14,-0.02]<sup>c</sup>** | -0.02 [-0.08, 0.04] | -0.09 [-0.14, -0.03]<sup>a</sup> | -0.04 [-0.10, 0.02] | -0.02 [-0.09, 0.04] | **-0.11 [-0.18, -0.04]<sup>b</sup>** |
| **Apathy** | **-0.18 [-0.30,-0.06]<sup>b</sup>** | -0.15 [-0.28, -0.02]<sup>a</sup> | -0.08 [-0.21, 0.04] | -0.16 [-0.29, -0.04]<sup>a</sup> | -0.01 [-0.14, 0.12] | -0.02 [-0.16, 0.11] |
| **Depression** | 0.01 [-0.12, 0.13] | 0.07 [-0.06, 0.21] | -0.08 [-0.21, 0.05] | 0.04 [-0.09, 0.17] | 0.11 [-0.03, 0.25] | -0.06 [-0.20, 0.09] |
| **Irritability** | -0.02 [-0.14, 0.10] | 0.00 [-0.13, 0.13] | -0.06 [-0.18, 0.06] | 0.02 [-0.11, 0.14] | 0.04 [-0.09, 0.17] | -0.20 [-0.33, -0.06]<sup>a</sup> |

Abbreviations: AD = Alzheimer's disease, MCI = mild cognitive impairment, MMSE = Mini-Mental State Examination, EF = executive
functions, NPI = neuropsychiatric inventory.
Data are standardized beta ( $\beta$ ) [95% confidence interval] corrected for sex, age, and education.
<sup>a</sup> $p < 0.05$ .
<sup>b</sup> $p < 0.05$ after correcting for false discovery rate.
<sup>c</sup> $p < 0.001$ after correcting for false discovery rate.

Next, age-, sex-, and education-corrected LMM assessing the associations between the presence of the three most common NPS and baseline cognitive performance showed that the presence of apathy was associated with worse MMSE scores in AD dementia (FDR-adjusted *p*<0.05). We found no associations between the presence of the three most common NPS and cognitive functioning at baseline in preclinical AD and MCI due to AD (all FDR-adjusted *p*>0.05, table 2).

Additional analyses examining associations between the remaining nine NPI-items and cognitive functioning showed that several other specific NPS were related to lower MMSE scores in AD dementia, nighttime behaviors were related to worse language performance in dementia, and the presence of hallucinations was associated with worse performance in attention in MCI due to AD (all FDR-adjusted *p*<0.05, table e-3).

### Associations between baseline NPS and cognitive functioning over time

Using LMMs adjusted for age, sex, and education, baseline NPI total scores were not associated with changes in MMSE scores or cognitive domains over time in our cohort of amyloid-*β* positive individuals ranging from preclinical AD to AD dementia (all FDR-adjusted *p*>0.05, table 3). With regard to specific NPS, baseline irritability was associated with less steep memory decline over time in individuals with preclinical AD at baseline (FDR-adjusted *p*<0.001). None of the baseline NPI-scores were associated with cognitive functioning over time in MCI due to AD and AD dementia (all FDR-adjusted *p*>0.05).

**Table 3.**
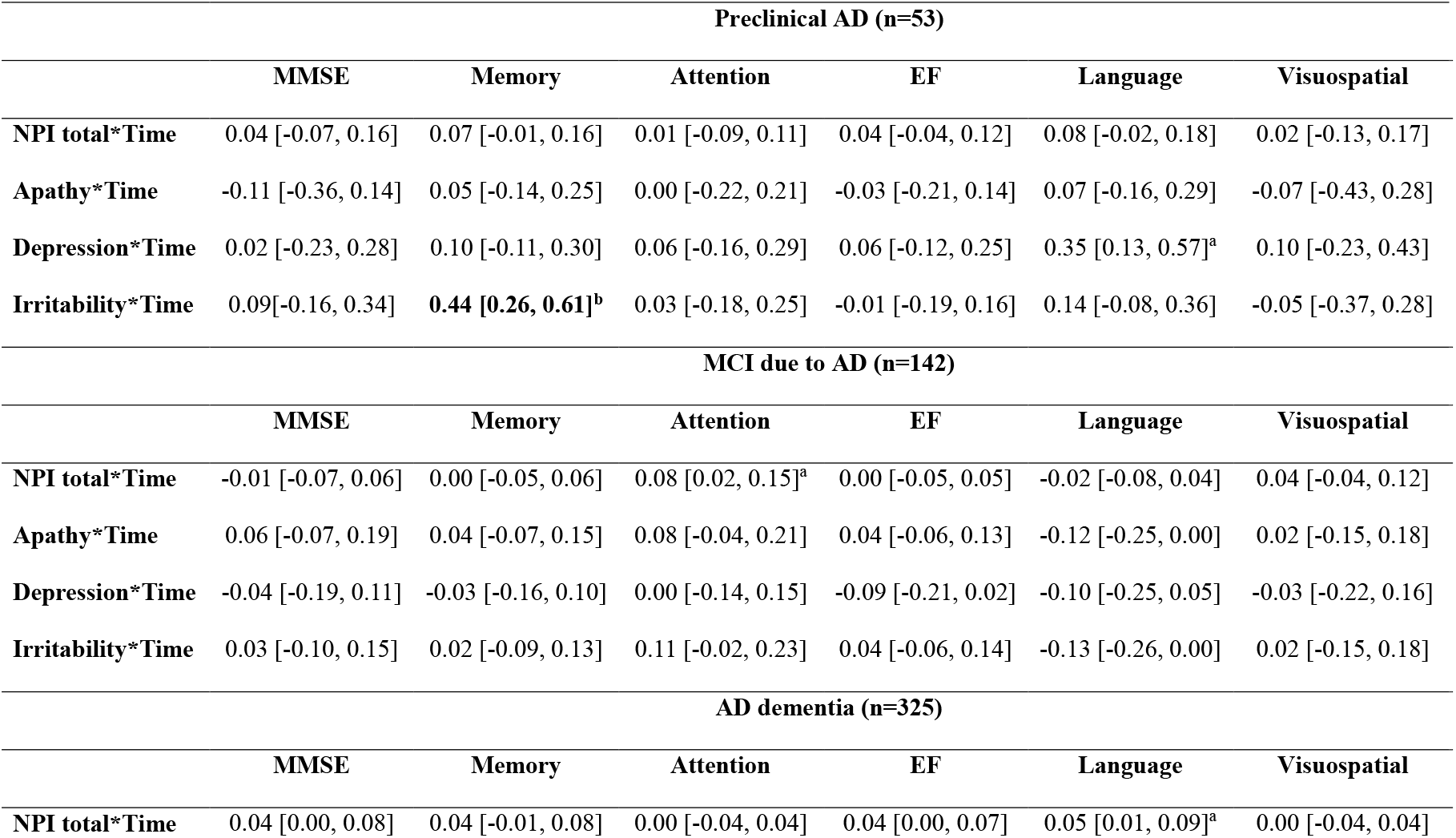
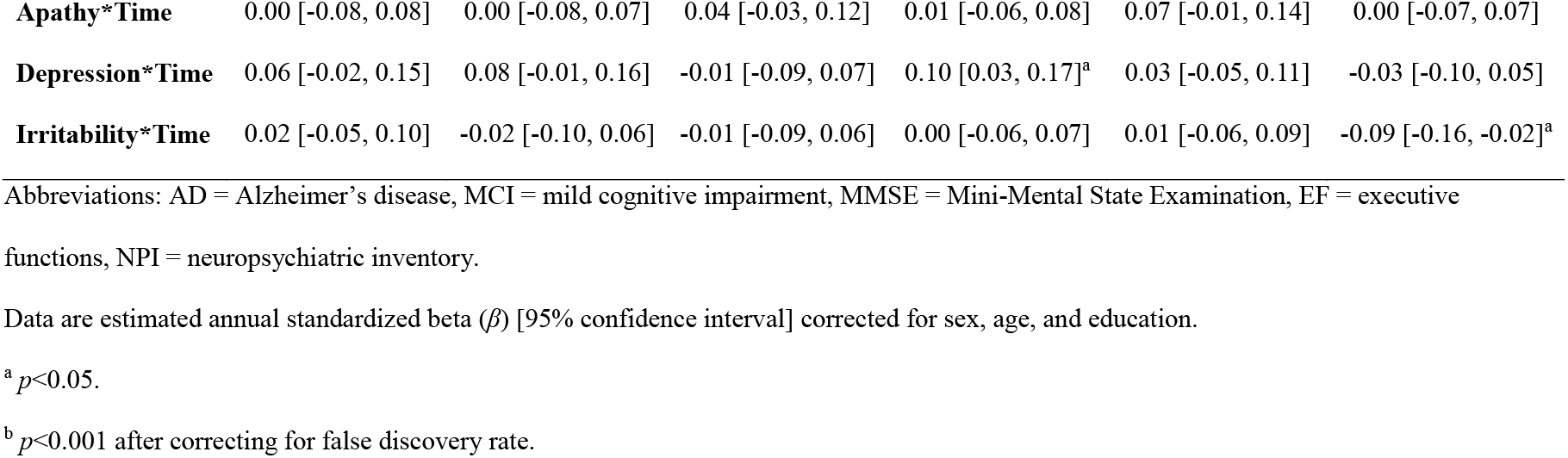
Associations between baseline NPS and cognitive decline over time according to clinical AD stage.

| **Preclinical AD (n=53)** |  |  |  |  |  |  |
| --- | --- | --- | --- | --- | --- | --- |
|  | **MMSE** | **Memory** | **Attention** | **EF** | **Language** | **Visuospatial** |
| **NPI total\*Time** | 0.04 [-0.07, 0.16] | 0.07 [-0.01, 0.16] | 0.01 [-0.09, 0.11] | 0.04 [-0.04, 0.12] | 0.08 [-0.02, 0.18] | 0.02 [-0.13, 0.17] |
| **Apathy\*Time** | -0.11 [-0.36, 0.14] | 0.05 [-0.14, 0.25] | 0.00 [-0.22, 0.21] | -0.03 [-0.21, 0.14] | 0.07 [-0.16, 0.29] | -0.07 [-0.43, 0.28] |
| **Depression\*Time** | 0.02 [-0.23, 0.28] | 0.10 [-0.11, 0.30] | 0.06 [-0.16, 0.29] | 0.06 [-0.12, 0.25] | 0.35 [0.13, 0.57]<sup>a</sup> | 0.10 [-0.23, 0.43] |
| **Irritability\*Time** | 0.09[-0.16, 0.34] | **0.44 [0.26, 0.61]<sup>b</sup>** | 0.03 [-0.18, 0.25] | -0.01 [-0.19, 0.16] | 0.14 [-0.08, 0.36] | -0.05 [-0.37, 0.28] |
| **MCI due to AD (n=142)** |  |  |  |  |  |  |
|  | **MMSE** | **Memory** | **Attention** | **EF** | **Language** | **Visuospatial** |
| **NPI total\*Time** | -0.01 [-0.07, 0.06] | 0.00 [-0.05, 0.06] | 0.08 [0.02, 0.15]<sup>a</sup> | 0.00 [-0.05, 0.05] | -0.02 [-0.08, 0.04] | 0.04 [-0.04, 0.12] |
| **Apathy\*Time** | 0.06 [-0.07, 0.19] | 0.04 [-0.07, 0.15] | 0.08 [-0.04, 0.21] | 0.04 [-0.06, 0.13] | -0.12 [-0.25, 0.00] | 0.02 [-0.15, 0.18] |
| **Depression\*Time** | -0.04 [-0.19, 0.11] | -0.03 [-0.16, 0.10] | 0.00 [-0.14, 0.15] | -0.09 [-0.21, 0.02] | -0.10 [-0.25, 0.05] | -0.03 [-0.22, 0.16] |
| **Irritability\*Time** | 0.03 [-0.10, 0.15] | 0.02 [-0.09, 0.13] | 0.11 [-0.02, 0.23] | 0.04 [-0.06, 0.14] | -0.13 [-0.26, 0.00] | 0.02 [-0.15, 0.18] |
| **AD dementia (n=325)** |  |  |  |  |  |  |
|  | **MMSE** | **Memory** | **Attention** | **EF** | **Language** | **Visuospatial** |
| **NPI total\*Time** | 0.04 [0.00, 0.08] | 0.04 [-0.01, 0.08] | 0.00 [-0.04, 0.04] | 0.04 [0.00, 0.07] | 0.05 [0.01, 0.09]<sup>a</sup> | 0.00 [-0.04, 0.04] |
| **Apathy\*Time** | 0.00 [-0.08, 0.08] | 0.00 [-0.08, 0.07] | 0.04 [-0.03, 0.12] | 0.01 [-0.06, 0.08] | 0.07 [-0.01, 0.14] | 0.00 [-0.07, 0.07] |
| **Depression\*Time** | 0.06 [-0.02, 0.15] | 0.08 [-0.01, 0.16] | -0.01 [-0.09, 0.07] | 0.10 [0.03, 0.17]<sup>a</sup> | 0.03 [-0.05, 0.11] | -0.03 [-0.10, 0.05] |
| **Irritability\*Time** | 0.02 [-0.05, 0.10] | -0.02 [-0.10, 0.06] | -0.01 [-0.09, 0.06] | 0.00 [-0.06, 0.07] | 0.01 [-0.06, 0.09] | -0.09 [-0.16, -0.02]<sup>a</sup> |

Abbreviations: AD = Alzheimer's disease, MCI = mild cognitive impairment, MMSE = Mini-Mental State Examination, EF = executive
functions, NPI = neuropsychiatric inventory.
Data are estimated annual standardized beta ( $\beta$ ) [95% confidence interval] corrected for sex, age, and education.
<sup>a</sup> $p < 0.05$ .
<sup>b</sup> $p < 0.001$ after correcting for false discovery rate.

We did not find associations across all clinical AD stages between the remaining nine NPI-items at baseline and cognitive performance over time (table e-4).

## DISCUSSION

The main findings in this amyloid-*β* positive sample are 1) high prevalence rates of NPS across all clinical AD stages, 2) a substantial heterogeneity in trajectories of NPS over time, 3) cross-sectional associations between the presence and severity of NPS and worse cognitive functioning in dementia, and 4) no clear associations between baseline NPS and performance on specific cognitive domains over time for all clinical AD stages.

NPS were prevalent across the entire clinical AD spectrum. Almost 90 percent of the patients with AD dementia showed at least one NPS, which is line with previous studies.^2, 21^ Furthermore, over 80 percent of the individuals in the pre-dementia AD stages exhibited at least one NPS, which is remarkably higher compared to prior studies.^6, 7^ Although NPS prevalence and severity was higher in AD dementia compared to pre-dementia AD stages, our findings suggest that NPS may precede cognitive impairment during the clinical course of AD. We did not find many differences in NPS prevalence and severity between individuals with preclinical AD and individuals with MCI due to AD, while some specific NPS were even more prevalent in preclinical AD compared to MCI due to AD. NPS that were common in the preclinical AD stage included affective symptoms, irritability, and nighttime behaviors and might be a psychological response to the initial cognitive decline experienced and might be a reason to visit the memory clinic.^22^ Prior studies have indeed shown higher NPS prevalence rates in pre-dementia memory clinic cohorts compared to population-based studies.^2, 22-24^ Our results indicate a different evolution over time for NPS compared to cognitive symptoms across the AD clinical spectrum. As expected, cognitive functioning showed a gradual decline that was most pronounced in the dementia stage.^25^ In contrast, the course of NPS showed a less coherent pattern with a relatively stable trends across all clinical AD stages, which is in line with prior research.^26, 27^ Moreover, we found substantial heterogeneity within individuals in their course of NPS compared to the intra-individual variation of the course of cognitive functioning over time. Although previous studies have also suggested large variability in NPS prevalence and evolution between and within patients,^28, 29^ the fluctuations in NPS observed in our study may also be due to the way NPS were measured. While cognitive functioning was assessed an extensive neuropsychological assessment covering five cognitive domains with at least two cognitive tests for each domain, NPS were measured using a single caregiver rating scale that can be affected by caregiver distress and recall bias.^3^ To obtain a better insight in fluctuations in NPS in AD, future studies could assess NPS on short time intervals using a combination of informant-based scales, clinician-rating instruments, and self-report measures, e.g. by using Ecological Momentary Assessments.^30^

At baseline, we found associations between the presence and severity of NPS and lower cognitive performance in patients with AD dementia that were most evident when looking at NPI total scores and MMSE performance. We found little evidence for a cross-sectional relationship between NPS and cognitive functioning in the pre-dementia AD stages. Cross-sectional relationships between NPS and cognitive deficits as measured with the MMSE have previously been reported in AD dementia,^31, 32^ and associations between NPS and performance on specific cognitive domains have also rarely been found in AD dementia.^5, 8, 33, 34^ Building further on this notion, we found no clear associations between baseline NPS and cognitive functioning over time across clinical stages. Although prior studies have yielded similar results in different cohorts of patients with AD dementia,^9, 33, 34^ our findings are in contrast to several other studies that have related the presence of NPS with accelerated cognitive decline in individuals with normal cognition,^11, 35^ MCI,^10^ and AD dementia.^4, 36^ This discrepancy in findings may be explained by the fact that the majority of these studies made use of the MMSE as global screening tool to assess cognition and the NPI total score to measure overall NPS burden. Although the MMSE is the most commonly used cognitive screening instrument for dementia, MMSE performance has been found to be correlated poorly with specific cognitive test scores and cognitive composite scores.^37^ Furthermore, assessing the presence and severity of NPS with the NPI total score might not fully capture the wide range of specific behavioral and psychological symptoms that may be present.

Previous studies have suggested that NPS are an integral part of AD and that the presence of NPS may be suggestive of underlying AD pathology.^38^ The studies described above that have previously examined the relationship between NPS and cognitive functioning have primarily included samples without AD-biomarker evidence. As a consequence, the presence of NPS in these samples may be suggestive of underlying AD pathology and has therefore been associated with cognitive decline. However, we already substantially increased the likelihood of underlying AD pathology as all individuals in our sample were amyloid-*β* positive. Consequently, the presence of NPS may have less predictive value in this sample of amyloid-*β* positive individuals.

Our findings provide useful information for the management of care for patients with AD. While one can expect a gradual decline of cognitive functioning over time, it appears difficult to predict the progression of NPS given the large differences between and within individuals despite group trajectories showing generally little change over time. These findings emphasize a patient-centered approach in the assessment and management of NPS across all clinical AD stages. Moreover, more future studies are needed that focus on identifying subgroups of individuals at risk for developing NPS.

Cognitive symptoms have been related to pathophysiologic and neurodegenerative processes in AD, with generally weaker associations with amyloid-*β* as compared to brain atrophy and tau pathology.^39^ Several theories have been proposed to explain the manifestation of NPS in AD.^3, 40^ While the *symptom hypothesis* states that NPS result from AD-related neuropathology that also contributes to cognitive impairment in AD, the *risk factor hypothesis* suggests that NPS arise from concurrent non-AD pathology, e.g. vascular depression.^40,41^ Recent studies have reported inconsistent associations between NPS and AD pathology,^42-44^ while providing some evidence for associations with non-AD biomarkers.^40, 45, 46^ Our findings suggested substantial fluctuations over time with no coherent pattern of decline or increase in NPS as the disease progresses leaving room open for other factors affecting NPS in AD. In addition to neurobiological causes, a variety of psychosocial factors have been proposed to play a role in the emergence and worsening of NPS in AD including unmet needs, stress among caregivers, and environmental triggers.^47^ Our findings show substantial fluctuations in NPS over time and no clear associations with cognitive symptoms suggesting that the *symptom hypothesis* alone cannot explain the emergence of NPS in AD.

Strengths of this study include the large well-defined sample of individuals who were all amyloid-*β* positive and underwent an extensive neuropsychological battery used to assess cognitive functioning. However, this study also has some limitations. First, although we took a unilateral perspective when examining the relationship between NPS and cognitive functioning, we acknowledge that cognitive impairments can also contribute to the presence and worsening of NPS in AD.^48^ Second, we examined a relatively young cohort of participants (M[SD] age=66.3[7.7]) who visited a tertiary memory clinic and may be characterized by a relative absence of age-related comorbidities. This may limit the generalizability of our findings to cohorts with older individuals with AD. Finally, we were not able to formally test NPS trajectories using LMM, as assumptions of normality and linearity were not met. This was caused by a substantial proportion of zero-scores on the NPI, as well as the way NPI-item-scores are calculated, i.e. by multiplying the severity score of 0 to 3 by the frequency score of 0 to 4 so that the values 5, 7, 10 and 11 cannot be observed.^49, 50^ Using symptom-specific instruments such as the Apathy Evaluation Scale (score range 18– 71), Cohen-Mansfield Agitation Inventory (score range 29–203), and Geriatric Depression Scale (score range 0–30) may not only help to fully characterize specific NPS, but also enables the use of statisical modelling due to a larger variation in potential scores compared to the NPI.

## CONCLUSIONS

NPS were prevalent in a well-defined amyloid-*β* positive sample ranging from normal cognition to dementia. We observed more heterogeneity in the course of NPS between and within individuals as compared to longitudinal cognitive functioning in AD. Furthermore, relating specific NPS to cognitive functioning, we found some cross-sectional associations in dementia but no longitudinal associations in any clinical AD stage. These findings suggest that NPS and cognitive symptoms are independent manifestations of AD that show a different evolution over the course of the disease.

## REFERENCES

1. McKhann GM, Knopman DS, Chertkow H, et al. The diagnosis of dementia due to Alzheimer’s disease: recommendations from the National Institute on Aging-Alzheimer’s Association workgroups on diagnostic guidelines for Alzheimer’s disease. Alzheimers Dement 2011;7:263–269.

2. Lyketsos CG, Lopez O, Jones B, Fitzpatrick AL, Breitner J, DeKosky S. Prevalence of neuropsychiatric symptoms in dementia and mild cognitive impairment: results from the cardiovascular health study. Jama 2002;288:1475–1483.

3. Geda YE, Schneider LS, Gitlin LN, et al. Neuropsychiatric symptoms in Alzheimer’s disease: past progress and anticipation of the future. Alzheimers Dement 2013;9:602–608.

4. Defrancesco M, Marksteiner J, Kemmler G, et al. Specific Neuropsychiatric Symptoms Are Associated with Faster Progression in Alzheimer’s Disease: Results of the Prospective Dementia Registry (PRODEM-Austria). J Alzheimers Dis 2020;73:125–133.

5. Koppel J, Goldberg TE, Gordon ML, et al. Relationships between behavioral syndromes and cognitive domains in Alzheimer disease: the impact of mood and psychosis. Am J Geriatr Psychiatry 2012;20:994–1000.

6. Pink A, Stokin GB, Bartley MM, et al. Neuropsychiatric symptoms, APOE epsilon4, and the risk of incident dementia: a population-based study. Neurology 2015;84:935–943.

7. Peters KR, Rockwood K, Black SE, et al. Characterizing neuropsychiatric symptoms in subjects referred to dementia clinics. Neurology 2006;66:523–528.

8. Ito T, Meguro K, Akanuma K, et al. Behavioral and psychological symptoms assessed with the BEHAVE-AD-FW are differentially associated with cognitive dysfunction in Alzheimer’s disease. J Clin Neurosci 2007;14:850–855.

9. Canevelli M, Adali N, Cantet C, et al. Impact of behavioral subsyndromes on cognitive decline in Alzheimer’s disease: data from the ICTUS study. J Neurol 2013;260:1859–1865.

10. David ND, Lin F, Porsteinsson AP. Trajectories of Neuropsychiatric Symptoms and Cognitive Decline in Mild Cognitive Impairment. Am J Geriatr Psychiatry 2016;24:70–80.

11. Burhanullah MH, Tschanz JT, Peters ME, et al. Neuropsychiatric Symptoms as Risk Factors for Cognitive Decline in Clinically Normal Older Adults: The Cache County Study. Am J Geriatr Psychiatry 2019:64–71.

12. Jack CR, Bennett DA, Blennow K, et al. NIA-AA Research Framework: Toward a biological definition of Alzheimer’s disease. Alzheimers Dement 2018;14:535–562.

13. van der Flier WM, Scheltens P. Amsterdam Dementia Cohort: Performing Research to Optimize Care. J Alzheimers Dis 2018;62:1091–1111.

14. Jessen F, Amariglio RE, van Boxtel M, et al. A conceptual framework for research on subjective cognitive decline in preclinical Alzheimer’s disease. Alzheimers Dement 2014;10:844–852.

15. Albert MS, DeKosky ST, Dickson D, et al. The diagnosis of mild cognitive impairment due to Alzheimer’s disease: recommendations from the National Institute on Aging-Alzheimer’s Association workgroups on diagnostic guidelines for Alzheimer’s disease. Alzheimers Dement 2011;7:270–279.

16. Duits FH, Teunissen CE, Bouwman FH, et al. The cerebrospinal fluid “Alzheimer profile”: easily said, but what does it mean? Alzheimers Dement 2014;10:713-723.e712.

17. Ossenkoppele R, Tolboom N, Foster-Dingley JC, et al. Longitudinal imaging of Alzheimer pathology using [11C]PIB, [18F]FDDNP and [18F]FDG PET. Eur J Nucl Med Mol Imaging 2012;39:990–1000.

18. Handels R, Jönsson L, Garcia-Ptacek S, Eriksdotter M, Wimo A. Controlling for selective dropout in longitudinal dementia data: Application to the SveDem registry. Alzheimers Dement 2020;16:789–796.

19. Cummings JL. The Neuropsychiatric Inventory: assessing psychopathology in dementia patients. Neurology 1997;48:S10–16.

20. Groot C, van Loenhoud AC, Barkhof F, et al. Differential effects of cognitive reserve and brain reserve on cognition in Alzheimer disease. Neurology 2018;90:e149–e156.

21. Zhao Q-F, Tan L, Wang H-F, et al. The prevalence of neuropsychiatric symptoms in Alzheimer’s disease: systematic review and meta-analysis. Journal of affective disorders 2016;190:264–271.

22. Sheikh F, Ismail Z, Mortby ME, et al. Prevalence of mild behavioral impairment in mild cognitive impairment and subjective cognitive decline, and its association with caregiver burden. Int Psychogeriatr 2018;30:233–244.

23. Siafarikas N, Selbaek G, Fladby T, Benth JŠ, Auning E, Aarsland D. Frequency and subgroups of neuropsychiatric symptoms in mild cognitive impairment and different stages of dementia in Alzheimer’s disease. Int Psychogeriatr 2018;30:103–113.

24. Geda YE, Roberts RO, Knopman DS, et al. Prevalence of neuropsychiatric symptoms in mild cognitive impairment and normal cognitive aging: population-based study. Arch Gen Psychiatry 2008;65:1193–1198.

25. Johnson DK, Storandt M, Morris JC, Galvin JE. Longitudinal study of the transition from healthy aging to Alzheimer disease. Arch Neurol 2009;66:1254–1259.

26. Brodaty H, Connors MH, Xu J, Woodward M, Ames D. The course of neuropsychiatric symptoms in dementia: a 3-year longitudinal study. J Am Med Dir Assoc 2015;16:380–387.

27. Vik-Mo AO, Giil LM, Borda MG, Ballard C, Aarsland D. The individual course of neuropsychiatric symptoms in people with Alzheimer’s and Lewy body dementia: 12-year longitudinal cohort study. Br J Psychiatry 2020;216:43–48.

28. Garre-Olmo J, Lopez-Pousa S, Vilalta-Franch J, de Gracia Blanco M, Vilarrasa AB. Grouping and trajectories of neuropsychiatric symptoms in patients with Alzheimer’s disease. Part II: two-year patient trajectories. J Alzheimers Dis 2010;22:1169–1180.

29. Tschanz JT, Corcoran CD, Schwartz S, et al. Progression of cognitive, functional, and neuropsychiatric symptom domains in a population cohort with Alzheimer dementia: the Cache County Dementia Progression study. Am J Geriatr Psychiatry 2011;19:532–542.

30. Niculescu L, Quirt H, Arora A, Green R, Ford B, Laboni A. Ecological momentary assessment for assessing depreession in advanced dementia: A pilot study. Am J Geriatr Psychiatry 2020;28:S120.

31. Spalletta G, Musicco M, Padovani A, et al. Neuropsychiatric symptoms and syndromes in a large cohort of newly diagnosed, untreated patients with Alzheimer disease. Am J Geriatr Psychiatry 2010;18:1026–1035.

32. Fernandez-Martinez M, Molano A, Castro JJ Zarranz J. Prevalence of neuropsychiatric symptoms in mild cognitive impairment and Alzheimer’s disease, and its relationship with cognitive impairment. Curr Alzheimer Res 2010;7:517–526.

33. Travis Seidl JN, Massman PJ. Cognitive and functional correlates of NPI-Q scores and symptom clusters in mildly demented Alzheimer patients. Alzheimer Dis Assoc Disord 2016;30:145–151.

34. Serra L, Perri R, Fadda L, et al. Relationship between cognitive impairment and behavioural disturbances in Alzheimer’s disease patients. Behav Neurol 2010;23:123–130.

35. Liew TM. Neuropsychiatric symptoms in cognitively normal older persons, and the association with Alzheimer’s and non-Alzheimer’s dementia. Alzheimers Res Ther 2020;12:1–14.

36. Rozum WJ, Cooley B, Vernon E, Matyi J, Tschanz JT. Neuropsychiatric symptoms in severe dementia: Associations with specific cognitive domains the Cache County Dementia Progression Study. Int J Geriatr Psychiatry 2019;34:1087–1094.

37. Jonaitis EM, Koscik RL, Clark LR, et al. Measuring longitudinal cognition: Individual tests versus composites. Alzheimers Dement 2019;11:74–84.

38. Geda YE, Krell-Roesch J, Sambuchi N, Michel BF. Neuropsychiatric symptoms and neuroimaging biomarkers in Alzheimer’s disease: “Which is the cart and which is the horse?”. Am J Geriatr Psychiatry 2017;25:694–696.

39. Ossenkoppele R, Smith R, Ohlsson T, et al. Associations between tau, Aβ, and cortical thickness with cognition in Alzheimer disease. Neurology 2019;92:e601–e612.

40. Peters ME, Lyketsos CG. Beyond memory: a focus on the other neuropsychiatric symptoms of dementia. Am J Geriatr Psychiatry 2015;23:115–118.

41. Alexopoulus GS, Meyers BS, Young RC, et al. ‘Vascular depression’ hypothesis. Arch Gen Psychiatry 1997;54:915–922.

42. Krell-Roesch J, Vassilaki M, Mielke MM, et al. Cortical β-amyloid burden, neuropsychiatric symptoms, and cognitive status: the Mayo Clinic Study of Aging. Transl Psychiatry 2019;9:1–8.

43. Malpas CB, Sharmin S, Kalincik T. The histopathological staging of tau, but not amyloid, corresponds to antemortem cognitive status, dementia stage, functional abilities and neuropsychiatric symptoms. Int J Neurosci 2020:1–10.

44. Banning LCP, Ramakers IHGB, Köhler S, et al. The association between biomarkers and neuropsychiatric symptoms across the Alzheimer’s disease spectrum. Am J Geriatr Psychiatry 2020;28:735–744.

45. Treiber KA, Lyketsos CG, Corcoran C, et al. Vascular factors and risk for neuropsychiatric symptoms in Alzheimer’s disease: the Cache County Study. Int Psychogeriatr 2008;20:538–553.

46. Bayram E, Shan G, Cummings JL. Associations between Comorbid TDP-43, Lewy Body Pathology, and Neuropsychiatric Symptoms in Alzheimer’s Disease. J Alzheimers Dis 2019;69:953–961.

47. Kales HC, Gitlin LN, Lyketsos CG. Assessment and management of behavioral and psychological symptoms of dementia. BMJ 2015;350:h369.

48. Rouch I, Padovan C, Boublay N, et al. Association between executive function and the evolution of behavioral disorders in Alzheimer’s disease. Int J Geriatr Psychiatry;35:1043–1050.

49. Saari T, Koivisto A, Hintsa T, Hänninen T, Hallikainen I. Psychometric Properties of the Neuropsychiatric Inventory: A Review. J Alzheimers Dis:1–15.

50. Hellton KH, Cummings J, Vik-Mo AO, et al. The truth behind the zeros: a new approach to principal component analysis of the Neuropsychiatric Inventory. Multivariate Behav Res 2020:1–16.

