## Supplemental Tables 1-4 for "Neuropsychiatric and cognitive symptoms across the Alzheimer’s disease clinical spectrum: Cross-sectional and longitudinal associations"

**Table e-1. Baseline neuropsychiatric inventory scores according to clinical AD stage.**

| | Preclinical AD (n=113) | | | MCI due to AD (n=321) | | | AD dementia (n=1,090) | | |
|---|---|---|---|---|---|---|---|---|---|
| | **Pre** | **Fre** | **Sev** | **Pre** | **Fre** | **Sev** | **Pre** | **Fre** | **Sev** |
| Apathy | 42.5% | 1.0 (1.3) | 0.7 (0.9) | 49.2% | 1.1 (1.2) | 0.8 (0.9) | 60.2%[a, e] | 1.5 (1.4)[a, b] | 1.1 (1.0)[a, b] |
| Irritability | 53.1% | 1.2 (1.2) | 0.9 (1.0) | 44.9% | 1.0 (1.2) | 0.8 (0.9) | 42.9% | 0.9 (1.2) | 0.7 (1.0) |
| Depression | 31.0% | 0.5 (0.9) | 0.4 (0.7) | 26.5% | 0.5 (1.0) | 0.4 (0.8) | 32.1% | 0.7 (1.1) | 0.5 (0.8) |
| Anxiety | 21.2% | 0.4 (0.9) | 0.3 (0.6) | 23.7% | 0.4 (0.8) | 0.4 (0.8) | 31.6%[b] | 0.7 (1.1)[c, d] | 0.5 (0.9)[d, e] |
| Eating behaviors | 21.2% | 0.4 ± 0.9 | 0.3 ± 0.7 | 20.1% | 0.4 ± 0.9 | 0.3 ± 0.7 | 31.4%[e] | 0.7 ± 1.1[b] | 0.5 ± 0.8[b] |
| Night-time behaviors | 30.1% | 0.7 ± 1.1 | 0.5 ± 0.8 | 20.1% | 0.4 ± 0.9 | 0.3 ± 0.7 | 20.0% | 0.5 ± 1.0 | 0.3 ± 0.7 |
| Agitation | 14.2 % | 0.3 (0.7) | 0.3 (0.7) | 15.3% | 0.3 (0.9) | 0.3 (0.7) | 16.2% | 0.3 (0.9) | 0.3 (0.7) |
| AMB | 8.0% | 0.2 ±0.7 | 0.1 ±0.4 | 12.1% | 0.3 ±0.8 | 0.2 ±0.5 | 18.9%[d, e] | 0.4 ± 1.0[d, e] | 0.3 ± 0.7[d, e] |
| Disinhibition | 9.7% | 0.2 (0.7) | 0.2 (0.6) | 13.7% | 0.2 ± 0.6 | 0.2 ± 0.6 | 12.4% | 0.2 ± 0.6 | 0.2 ± 0.6 |
| Delusions | 3.5% | 0.1 ±0.4 | 0.1 ±0.4 | 4.7% | 0.1 ±0.5 | 0.1 ±0.3 | 9.3%[d, e] | 0.2 ± 0.7[e] | 0.2 ± 0.5[e] |
| Euphoria | 4.4% | 0.1 (0.3) | 0.1 (0.4) | 5.3% | 0.1 (0.5) | 0.1 (0.4) | 6.0% | 0.1 (0.4) | 0.1 (0.3) |
| Hallucinations | 5.3% | 0.1 (0.6) | 0.1 (1.1) | 1.2% | 0.0 ± 0.2 | 0.0 ± 0.2 | 4.6%[e] | 0.1 ± 0.4 | 0.1 ± 0.3 |
| Overall | 81.4% | 5.1 ± 4.6 | 3.8 ± 3.6 | 81.2% | 4.8 ± 4.5 | 3.8 ± 3.6 | 88.7%[e] | 6.3 ± 5.1[b, d] | 4.8 ± 3.9[b,d] |

*Notes.* AD = Alzheimer's disease, MCI = mild cognitive impairment, Pre = prevalence (NPI frequency × severity scores ≥1), Fre = NPI frequency score (range 0–4), Sev = NPI severity score (range 0–3), AMB = aberrant motor behaviors. Differences in prevalence rates tested using Chi-Square Tests, differences in frequency and severity scores tested using Kruskal-Wallis Tests.

[a] preclinical AD < AD dementia, $p<0.001$ after correcting for false discovery rate.

[b] MCI due to AD < AD dementia, $p<0.001$ after correcting for false discovery rate.

[c] MCI due to AD < AD dementia, $p<0.01$ after correcting for false discovery rate.

[d] preclinical AD < AD dementia, $p<0.05$ after correcting for false discovery rate.

[e] MCI due to AD < AD dementia, $p<0.05$ after correcting for false discovery rate.

**Table e-2. Within-subject and between-subject variances of cognitive and neuropsychiatric measures over time according to clinical AD stage.**

| | **Preclinical AD at baseline** | | **MCI due to AD at baseline** | | **AD dementia at baseline** | |
|---|---|---|---|---|---|---|
| | *BS variance (% explained)* | *WS variance (% explained)* | *BS variance (% explained)* | *WS variance (% explained)* | *BS variance (% explained)* | *WS variance (% explained)* |
| **Cognitive measures** | | | | | | |
| **MMSE** | 1.42 (52.40%) | 1.29 (47.60%) | 3.51 (47.37%) | 3.90 (52.63%) | 14.55 (54.29%) | 12.25 (45.71%) |
| **Memory** | 0.64 (64.65%) | 0.35 (35.35%) | 2.09 (59.71%) | 1.41 (40.29%) | 2.75 (60.04%) | 1.83 (36.96%) |
| **Attention** | 0.48 (64.86%) | 0.26 (35.14%) | 1.07 (56.91%) | 0.81 (43.09%) | 10.73 (62.60%) | 6.41 (37.40%) |
| **Executive functions** | 0.61 (78.21%) | 0.17 (21.79%) | 0.87 (74.36%) | 0.30 (25.64%) | 2.14 (70.86%) | 0.88 (29.14%) |
| **Language** | 0.19 (61.29%) | 0.12 (38.71%) | 0.36 (61.02%) | 0.23 (38.98%) | 1.86 (64.36%) | 1.03 (35.64%) |
| **Visuospatial** | 0.08 (17.78%) | 0.37 (82.22%) | 0.49 (37.12%) | 0.83 (62.88%) | 11.72 (73.76%) | 4.17 (26.24%) |
| **Average % explained** | 56.53% | 43.47% | 56.08% | 43.90% | 64.32% | 35.18% |
| **NPS measures** | | | | | | |
| **NPI total score** | 36.11 (46.16%) | 42.12 (56.84%) | 29.70 (37.85%) | 48.77 (62.15%) | 41.03 (37.25%) | 69.12 (62.75%) |
| **NPI-apathy** | 0.84 (13.93%) | 5.19 (86.07%) | 1.40 (22.29%) | 4.88 (77.71%) | 2.24 (20.25%) | 8.82 (79.75%) |

| | | | | | | |
|---|---|---|---|---|---|---|
| **NPI-depression** | 0.76 (25.25%) | 2.25 (74.75%) | 1.45 (31.94%) | 3.09 (68.06%) | 1.40 (29.60%) | 3.33 (70.40%) |
| **NPI-irritability** | 2.23 (27.23%) | 5.96 (72.77%) | 1.43 (23.40%) | 4.68 (76.60%) | 2.16 (30.00%) | 5.04 (70.00%) |
| **Average % explained** | 28.14% | 72.61% | 28.87% | 71.13% | 29.28% | 70.73% |

Abbreviations: BS = between-subject, WS = within-subject, AD = Alzheimer's disease, MCI = mild cognitive impairment, MMSE = Mini-Mental State Examination.

**Table e-3. Associations between baseline NPS and concurrent cognitive performance according to clinical AD stage.**

| | Preclinical AD (n=113) | | | | | |
|---|---|---|---|---|---|---|
| | **MMSE** | **Memory** | **Attention** | **EF** | **Language** | **Visuospatial** |
| **Agitation** | 0.22 [-0.25, 0.69] | 0.43 [-0.08, 0.93] | -0.02 [-0.51, 0.48] | -0.20 [-0.68, 0.27] | -0.35 [-0.86, 0.16] | 0.35 [-0.09, 0.80] |
| **AMB** | 0.01 [-0.61, 0.63] | 0.31 [-0.37, 0.99] | 0.32 [-0.32, 0.96] | -0.20 [-0.81, 0.42] | 0.11 [-0.59, 0.80] | 0.30 [-0.31, 0.92] |
| **Anxiety** | 0.12 [-0.41, 0.65] | 0.05 [-0.46, 0.55] | 0.36 [-0.15, 0.86] | 0.14 [-0.32, 0.60] | 0.05 [-0.47, 0.58] | 0.22 [-1.03, 1.46] |
| **Delusions** | 1.19 [0.06, 2.32][a] | 0.47 [-0.67, 1.61] | 1.22 [0.11, 2.33][a] | 1.01 [-0.03, 2.06] | -0.45 [-1.62, 0.72] | 1.25 [-0.90, 3.40] |
| **Disinhibition** | -0.45 [-0.97, 0.07] | -0.12 [-0.72, 0.47] | -0.21 [-0.78, 0.36] | -0.25 [-0.79, 0.29] | 0.02 [-0.58, 0.62] | 0.25 [-0.27, 0.78] |
| **Eating behaviors** | 0.21 [-0.22, 0.63] | 0.05 [-0.39, 0.49] | 0.05 [-0.38, 0.49] | -0.09 [-0.50, 0.32] | -0.13 [-0.59, 0.32] | 0.57 [0.14, 1.01][a] |
| **Euphoria** | 0.30 [-0.46, 1.06] | 0.35 [-0.47, 1.18] | 0.40 [-0.39, 1.20] | 0.42 [-0.34, 1.18] | -0.27 [-1.10, 0.56] | 0.15 [-0.54, 0.85] |
| **Hallucinations** | 0.36 [-0.52, 1.23] | 0.22 [-0.71, 1.16] | 0.24 [-0.67, 1.15] | -0.24 [-1.11, 0.62] | 0.19 [-0.75, 1.14] | 0.71 [-0.12, 1.55] |
| **Night-time behaviors** | 0.09 [-0.26, 0.45] | 0.42 [0.04, 0.79] | 0.25 [-0.11, 0.62] | 0.39 [0.04, 0.73][a] | 0.17 [-0.21, 0.55] | -0.15 [-0.50, 0.20] |
| | **MCI due to AD (n=321)** | | | | | |
| | **MMSE** | **Memory** | **Attention** | **EF** | **Language** | **Visuospatial** |
| **Agitation** | 0.10 [-0.16, 0.36] | 0.11 [-0.17, 0.40] | -0.23 [-0.51, 0.05] | -0.21 [-0.47, 0.06] | -0.02 [-0.32, 0.28] | -0.01 [-0.30, 0.28] |

| | | | | | | |
|---|---|---|---|---|---|---|
| **AMB** | 0.01 [-0.30, 0.31] | -0.10 [-0.44, 0.23] | -0.36 [-0.70, -0.03][a] | 0.00 [-0.32, 0.31] | -0.19 [-0.54, 0.16] | 0.17 [-0.19, 0.53] |
| **Anxiety** | 0.13 [-0.09, 0.35] | 0.00 [-0.24, 0.25] | -0.05 [-0.29, 0.19] | -0.02 [-0.25, 0.21] | -0.10 [-0.35, 0.16] | 0.04 [-0.21, 0.29] |
| **Delusions** | -0.12 [-0.57, 0.32] | -0.51 [-0.99, -0.04][a] | -0.24 [-0.72, 0.24] | 0.00 [-0.45, 0.46] | 0.20 [-0.31, 0.71] | 0.18 [-0.30, 0.65] |
| **Disinhibition** | 0.13 [-0.14, 0.41] | 0.41 [0.11, 0.71][a] | -0.20 [-0.50, 0.10] | -0.03 [-0.31, 0.25] | -0.17 [-0.49, 0.15] | 0.17 [-0.14, 0.48] |
| **Eating behaviors** | 0.00 [-0.24, 0.24] | -0.04 [-0.30, 0.22] | -0.21 [-0.47, 0.05] | -0.23 [-0.47, 0.02] | -0.41 [-0.69, -0.14][a] | -0.10 [-0.38, 0.18] |
| **Euphoria** | 0.34 [-0.07, 0.74] | 0.20 [-0.23, 0.64] | 0.08 [-0.35, 0.52] | 0.08 [-0.33, 0.50] | 0.20 [-0.26, 0.66] | -0.36 [-0.80, 0.08] |
| **Hallucinations** | -0.29 [-1.21, 0.62] | -0.62 [-1.55, 0.31] | **-1.73 [-2.68, -0.79][b]** | -1.16 [-2.04, -0.29][a] | -0.28 [-1.28, 0.72] | -0.58 [-1.69, 0.53] |
| **Night-time behaviors** | 0.25 [-0.02, 0.48] | 0.11 [-0.14, 0.37] | -0.03 [-0.28, 0.22] | 0.06 [-0.18, 0.30] | 0.16 [-0.11, 0.43] | 0.00 [-0.27, 0.27] |
| | | | **AD dementia (n=1,090)** | | | |
| | **MMSE** | **Memory** | **Attention** | **EF** | **Language** | **Visuospatial** |
| **Agitation** | **-0.27 [-0.43, -0.11][b]** | -0.01 [-0.19, 0.17] | -0.11 [-0.28, 0.05] | -0.17 [-0.34, 0.00] | -0.08 [-0.25, 0.10] | -0.09 [-0.28, 0.10] |
| **AMB** | **-0.28 [-0.43, -0.13][b]** | -0.16 [-0.33, 0.01] | -0.22 [-0.37, 0.06] | -0.09 [-0.25, 0.07] | -0.05 [-0.21, 0.12] | -0.20 [-0.38, -0.02][a] |
| **Anxiety** | -0.13 [-0.26, 0.00] | -0.08 [-0.22, 0.06] | -0.13 [-0.26, 0.00] | -0.03 [-0.16, 0.11] | -0.01 [-0.15, 0.13] | -0.20 [-0.34, -0.05][a] |
| **Delusions** | -0.16 [-0.38, 0.06] | -0.15 [-0.39, 0.10] | -0.26 [-0.49, -0.04][a] | -0.18 [-0.40, 0.05] | -0.13 [-0.37, 0.10] | -0.41 [-0.65, -0.16][a] |
| **Disinhibition** | 0.02 [-0.16, 0.19] | 0.11 [-0.08, 0.30] | 0.08 [-0.10, 0.27] | -0.03 [-0.21, 0.16] | 0.03 [-0.17, 0.22] | -0.07 [-0.27, 0.13] |

| | | | | | | |
|---|---|---|---|---|---|---|
| **Eating behaviors** | -0.07 [-0.20, 0.05] | 0.01 [-0.13, 0.15] | -0.08 [-0.21, 0.05] | -0.08 [-0.22, 0.05] | -0.03 [-0.17, 0.11] | -0.06 [-0.20, 0.09] |
| **Euphoria** | **-0.27 [-0.41, -0.12][b]** | -0.17 [-0.33, 0.00] | -0.17 [-0.33, -0.02][a] | -0.07 [-0.23, 0.08] | -0.06 [-0.22, 0.10] | -0.19 [-0.37, -0.02][a] |
| **Hallucinations** | -0.41 [-0.70, -0.12][a] | -0.02 [-0.36, 0.31] | -0.35 [-0.65, -0.05][a] | -0.37 [-0.68, -0.06][a] | -0.09 [-0.40, 0.22] | -0.29 [-0.66, 0.07] |
| **Night-time behaviors** | -0.13 [-0.27, 0.01] | 0.07 [-0.09, 0.23] | -0.05 [-0.20, 0.10] | 0.01 [-0.14, 0.16] | **-0.29[-0.44, -0.13][a]** | -0.07 [-0.07, 0.09] |

Abbreviations: AD = Alzheimer's disease, MCI = mild cognitive impairment, MMSE = Mini-mental state examination, EF = executive functions, AMB = aberrant motor behaviors.

Data are standardized estimated annual $\beta$ [95% confidence interval] corrected for sex, age, and education.

[a] $p<0.05$.

[b] $p<0.05$ after correcting for false discovery rate.

**Table e-4. Associations between baseline NPS and cognitive decline over time according to clinical AD stage.**

| | Preclinical AD (n=53) | | | | | |
|---|---|---|---|---|---|---|
| | **MMSE** | **Memory** | **Attention** | **EF** | **Language** | **Visuospatial** |
| **Agitation*Time** | 0.11 [-0.24, 0.46] | -0.03 [-0.30, 0.24] | -0.03 [-0.33, 0.27] | -0.23 [-0.47, 0.01] | 0.00 [-0.31, 0.31] | 0.03 [-0.40, 0.46] |
| **AMB*Time** | 0.28 [-0.12, 0.68] | 0.24 [-0.09, 0.58] | -0.19 [-0.55, 0.17] | -0.17 [-0.47, 0.13] | 0.30 [-0.07, 0.67] | 0.05 [-0.45, 0.56] |
| **Anxiety*Time** | 0.01 [-0.60, 0.63] | -0.18 [-0.66, 0.30] | 0.74 [0.25, 1.24][a] | 0.29 [-0.15, 0.73] | 0.05 [-0.49, 0.59] | -0.13 [-2.05, 1.79] |
| **Delusions*Time** | 0.57 [-0.90, 2.03] | 0.12 [-0.97, 1.22] | 0.06 [-1.16, 1.28] | 0.14 [-0.86, 1.15] | -0.62 [-1.87, 0.62] | 1.96 [-1.67, 5.59] |
| **Disinhibition* *Time** | 0.12 [-0.22, 0.47] | 0.05 [-0.23, 0.32] | -0.12 [-0.43, 0.18] | -0.19 [-0.44, 0.06] | 0.14 [-0.18, 0.45] | 0.03 [-0.40, 0.46] |
| **Eating behaviors*Time** | 0.25 [-0.15, 0.64] | 0.17 [-0.15, 0.48] | 0.04 [-0.30, 0.39] | 0.19 [-0.10, 0.47] | 0.00 [-0.36, 0.35] | 0.02 [-0.47, 0.51] |
| **Euphoria*Time** | -0.08 [-0.61, 0.45] | -0.19 [-0.59, 0.21] | -0.03 [-0.47, 0.42] | -0.12 [-0.49, 0.25] | -0.24 [-0.70, 0.21] | -0.02 [-0.66, 0.62] |
| **Hallucinations* Time** | 0.13 [-0.43, 0.68] | 0.01 [-0.42, 0.44] | -0.03 [-0.50, 0.45] | 0.50 [0.12, 0.88][a] | 0.15 [-0.33, 0.64] | -0.11 [-0.79, 0.58] |
| **Night-time behaviors*Time** | 0.02 [-0.23, 0.28] | 0.18 [-0.02, 0.38] | 0.14 [-0.08, 0.36] | 0.12 [-0.06, 0.30] | 0.12 [-0.11, 0.35] | 0.13 [-0.20, 0.47] |
| | **MCI due to AD (n=142)** | | | | | |
| | **MMSE** | **Memory** | **Attention** | **EF** | **Language** | **Visuospatial** |

| | | | | | | |
|---|---|---|---|---|---|---|
| **Agitation*Time** | 0.03 [-0.13, 0.20] | 0.05 [-0.10, 0.19] | 0.00 [-0.16, 0.16] | 0.03 [-0.09, 0.15] | 0.07 [-0.09, 0.22] | -0.03 [-0.24, 0.18] |
| **AMB*Time** | 0.08 [-0.17, 0.32] | 0.15 [-0.05, 0.36] | -0.09 [-0.33, 0.15] | 0.11 [-0.07, 0.29] | 0.05 [-0.18, 0.28] | 0.07 [-025, 0.38] |
| **Anxiety*Time** | 0.14 [-0.02, 0.30] | -0.04 [-0.17, 0.10] | 0.05 [-0.11, 0.20] | 0.08 [-0.04, 0.20] | 0.03 [-0.13, 0.18] | 0.10 [-0.09, 0.29] |
| **Delusions*Time** | 0.05 [-0.30, 0.40] | 0.06 [-0.23, 0.35] | 0.06 [-0.28, 0.39] | -0.04 [-0.30, 0.22] | -0.03 [-0.37, 0.30] | 0.03 [-0.39, 0.45] |
| **Disinhibition* Time** | 0.01 [-0.19, 0.22] | 0.14 [-0.04, 0.33] | 0.07 [-0.14, 0.27] | 0.11 [-0.04, 0.27] | 0.15 [-0.06, 0.35] | 0.08 [-0.17, 0.33] |
| **Eating behaviors*Time** | 0.01 [-0.16, 0.17] | -0.04 [-0.18, 0.10] | 0.00 [-0.16, 0.16] | -0.07 [-0.19, 0.06] | -0.25 [-0.41, -0.08][a] | -0.10 [-0.30, 0.11] |
| **Euphoria*Time** | -0.20 [-0.42, 0.02] | -0.14 [-0.32, 0.04] | -0.27 [-0.48, -0.06][a] | -0.26 [-0.42, -0.10][a] | -0.07 [-0.28, 0.14] | -0.23 [-0.49, 0.04] |
| **Hallucinations* Time** | N/A[#] | N/A[#] | N/A[#] | N/A[#] | N/A[#] | N/A[#] |
| **Night-time behaviors*Time** | 0.14 [-0.01, 0.30] | 0.06 [-0.07, 0.19] | 0.02 [-0.13, 0.17] | -0.09 [-0.21, 0.02] | 0.07 [-0.09, 0.22] | -0.09 [-0.28, 0.10] |
| | | | **AD dementia (n=325)** | | | |
| | **MMSE** | **Memory** | **Attention** | **EF** | **Language** | **Visuospatial** |
| **Agitation*Time** | -0.07 [-0.18, 0.04] | 0.00 [-0.11, 0.12] | -0.04 [-0.15, 0.07] | 0.00 [-0.10, 0.10] | 0.07 [-0.04, 0.18] | -0.08 [-0.18, 0.02] |
| **AMB*Time** | -0.03 [-0.14, 0.07] | -0.02 [-0.13, 0.08] | -0.15 [-0.25, -0.05][a] | -0.09 [-0.18, 0.00] | 0.05 [-0.06, 0.15] | -0.02 [-0.12, 0.08] |

| | | | | | | |
|---|---|---|---|---|---|---|
| **Anxiety*Time** | 0.02 [-0.06, 0.10] | -0.02 [-0.10, 0.07] | -0.06 [-0.14, 0.02] | 0.01 [-0.06, 0.08] | 0.03 [-0.05, 0.11] | -0.01 [-0.08, 0.07] |
| **Delusions*Time** | 0.12 [-0.07, 0.32] | -0.14 [-0.35, 0.06] | -0.14 [-0.33, 0.06] | -0.03 [-0.20, 0.14] | -0.01 [-0.21, 0.19] | -0.24 [-0.41, -0.07] |
| **Disinhibition*Time** | 0.09 [-0.03, 0.21] | -0.09 [-0.22, 0.04] | -0.02 [-0.14, 0.10] | 0.01 [-0.10, 0.12] | 0.13 [0.01, 0.25][a] | 0.09 [-0.02, 0.20] |
| **Eating behaviors*Time** | 0.01 [-0.07, 0.08] | 0.00 [-0.08, 0.08] | 0.00 [-0.08, 0.07] | -0.01 [-0.08, 0.06] | 0.05 [-0.03, 0.13] | 0.04 [-0.03, 0.11] |
| **Euphoria*Time** | -0.06 [-0.22, 0.11] | -0.24 [-0.44, -0.05][a] | -0.09 [-0.28, 0.09] | -0.19 [-0.35, -0.03][a] | 0.12 [-0.06, 0.31] | -0.02 [-0.17, 0.14] |
| **Hallucinations*Time** | -0.09 [-0.29, 0.10] | 0.10 [-0.11, 0.32] | 0.05 [-0.15, 0.26] | 0.09 [-0.09, 0.28] | 0.04 [-0.16, 0.24] | 0.06 [-0.13, 0.25] |
| **Night-time behaviors*Time** | 0.03 [-0.06, 0.12] | -0.02 [-0.11, 0.07] | 0.12 [0.03, 0.20][a] | 0.06 [-0.02, 0.14] | 0.04 [-0.05, 0.13] | 0.01 [-0.07, 0.09] |

Abbreviations: AD = Alzheimer's disease, MCI = mild cognitive impairment, MMSE = Mini-mental state examination, EF = executive functions, AMB = aberrant motor behaviors.

Data are standardized estimated annual $\beta$ [95% confidence interval] corrected for sex, age, and education.

[a] $p<0.05$.

[b] $p<0.05$ after correcting for false discovery rate.

[#] No patients with MCI and hallucinations at baseline had follow-up assessments available.
